# Cognitive profile accurately predicts underlying pathology in Corticobasal Syndrome

**DOI:** 10.64898/2025.12.17.25342469

**Authors:** Steven Smithies, Amir Ebneabbasi, Riccardo Conci, Alexander Murley, George Savulich, Katherine Stockton, Matthew Rouse, Michelle Naessens, Maura Malpetti, Timothy Rittman, Kieren Allinson, Annelies Quaegebeur, James Rowe, Negin Holland

## Abstract

**Background/Objectives:** Corticobasal syndrome (CBS) shows weak clinicopathological correlation, with only half of cases attributable to corticobasal degeneration (CBD). Common alternative aetiologies include Alzheimer’s disease (AD) and progressive supranuclear palsy (PSP). Emerging disease modifying therapies are likely to require accurate antemortem differentiation. While plasma biomarkers have potential, clinical/cognitive tools such as the Addenbrooke’s Cognitive Examination-Revised (ACE-R), Cambridge Behavioural Inventory-Revised (CBI-R), and PSP Rating Scale (PSP-RS) may provide a cost-effective and scalable alternative.

**Methods:** We retrospectively analysed 226 people diagnosed with CBS between 1997–2024, of whom 72 had post-mortem neuropathological assessment. We compared ACE-R, CBI-R, and PSP-RS subdomain scores across pathologies and trained predictive models to differentiate them. In a subset of 52 patients, we also evaluated plasma biomarkers (Neurofilament Light Chain, pTau217, Aβ42/Aβ40 ratio) as indicators of AD pathology.

**Results:** People with CBS with AD pathology showed significantly worse scores in ACE-R memory (p=0.002), visuospatial (p=0.002) and attention (p=0.012), and CBI-R memory & orientation (p=0.022) domains. Principal component analysis identified a ‘global cognition’ factor (PC2), with higher scores in AD cases (i.e: worse cognition; p=<0.001). Multiple predictive models trained on ACE-R/CBI-R subdomain scores predicted AD pathology with 84.9-85.6% accuracy (AUC 0.874-0.926). Plasma biomarkers failed to reliably distinguish the dominant underlying pathology, in our cohort.

**Conclusions:** Neurocognitive assessments offer a rapid, inexpensive, and scalable method to identify AD pathology in CBS, outperforming current plasma biomarkers. These tools may facilitate targeted trial recruitment and clinical management decisions, while additional development of blood biomarkers is required in the context of corticobasal syndrome.

## Introduction

The Corticobasal Syndrome (CBS) is a clinical construct characterised by a progressive, asymmetric limb rigidity, akinesia, dystonia or myoclonus as well as apraxia, cortical sensory deficit and alien limb phenomenon (1). Underlying aetiologies include Corticobasal Degeneration (CBD, ∼50%), Alzheimer’s disease (AD, 30-40%), Progressive Supranuclear Palsy (PSP, 10-30%), α-Synucleinopathies such as Lewy-body disease, and Frontotemporal Lobar degeneration - TDP43 (FTLD-TDP43) (2–5). CBD is a distinct neuropathological 4-repeat tauopathy with characteristic tau-positive neuronal and astroglial lesions (6). CBD pathology can present with syndromes other than CBS, such as PSP or behavioural variant frontotemporal dementia (bvFTD) (2,3,5,7,8), but here we focus on people with CBS

The weak clinicopathological correlation of CBS poses challenges for diagnosis, prognostication, and recruitment into pathology-specific therapeutic trials (2). Although post-mortem examination remains the diagnostic gold standard, most clinicopathological studies are underpowered (3–5,7,9–14), while larger clinical studies often lack pathological confirmation (15–18). As disease-modifying therapies targeting specific pathologies emerge, accurate antemortem differentiation becomes increasingly critical - not only for trial stratification but also for interpreting outcomes in basket trials.

Blood-based biomarkers such as plasma pTau217, Aβ42:Aβ40 ratio, and neurofilament light chain (NFL) show promise as markers of AD pathology in the context of amnestic and non-amnestic cognitive syndromes and normal ageing (3,19). For example, higher plasma pTau217 levels and lower Aβ42:Aβ40 ratios have been validated as markers of AD pathology with high accuracy (20–22). Neurofilament Light Chain (NFL) is a less specific marker of neurodegeneration (23–25). However, these tests are not universally available and cannot simply be assumed to differentiate the aetiology of CBS.

Clinical tests and scales offer a scalable, low-cost alternative. Examples include the Addenbrooke’s Cognitive Examination-Revised (ACE-R), Cambridge Behavioural Inventory-Revised (CBI-R), and PSP Rating Scale (PSP-RS). Such cognitive and behavioural profiles combined with machine learning methods, can enhance pathological prediction (15,16).

The ACE has been validated for diagnosing dementia (26), including in CBS (27,28), with differing patterns of performance distinguishing AD from other neurodegenerative diseases (29). Similar evidence exists for carer endorsements on the CBI (16).

Here we test the hypothesis that clinical scoring systems can accurately differentiate underlying pathology in CBS, particularly AD vs non-AD pathologies. We compare this performance to blood biomarkers. Although singular clinical features do not accurately differentiate underlying pathology (2), combinations of features may do so using simple machine learning.

## Methods

### Participants

We identified 226 patients who received a clinical diagnosis of Corticobasal Syndrome (CBS) at their initial clinical visit to either the Memory Clinic or the Disorders of Movement and Cognition Clinics at Cambridge University Hospitals and the Cambridge Centre for Parkinson-plus between 1997 and 2024. Of these, 72 patients had post-mortem neuropathological examination. All 72 met the 2013 Armstrong criteria for CBS during life (1).

### Clinical Assessment

All participants underwent a clinical assessment with history, examination and the Addenbrooke’s Cognitive Examination (ACE or ACE-revised), the informant-rated Cambridge Behavioural Inventory (CBI or CBI-revised), and the Progressive Supranuclear Palsy Rating Scale (PSP-RS, only available from 2007 onwards). From 2006, the revised version of the ACE (ACE-R) was introduced; and from 2008 the revised CBI-R was introduced. Earlier ACE and CBI assessments were retrospectively rescored based on their original answers to align with the revised versions, ensuring consistency across the dataset.

To standardise comparisons, all scores were converted to a proportion of the total possible score (range: 0–1). Since higher ACE scores indicate better cognitive performance, ACE scores were inverted to reflect the proportion of points lost, aligning directionality with the CBI and PSP-RS (where higher scores reflect greater impairment). Scores from the initial clinical assessment at first visit were used in the analysis.

### Plasma biomarkers

A subset of 52 patients also had plasma biomarkers available (pTau217, Neurofilament Light Chain (NFL), Aβ42 and Aβ40 levels). The details of sample collection and analysis are outlined in (30). Cut-off values for each biomarker were as follows: pTau217: 3 range approach with lower (95% sensitivity, <0.4pg/ml) & upper (95% specificity, 0.63pg/ml) cutoffs (20); NFL: >38.04pg/ml (23); Aβ42:Aβ40 ratio: <0.12 (22,31).

### Pathological diagnosis

72 patients underwent post-mortem neuropathological examination. Primary and co-pathologies were registered following assessment of hyperphosphorylated tau, amyloid beta, alpha-synuclein and TDP43 pathology in key regions of the left hemisphere. Significant AD co-pathology was defined as intermediate and high level of AD neuropathological change as per the National Institute on Aging–Alzheimer’s Association guideline (32). All 72 cases met the 2013 Armstrong criteria for CBS during life (1).

### Statistical analysis

All analyses were performed in RStudio (version 4.5.1). The following steps were undertaken:

1. **Group differences in clinical characteristics:** For participants with pathological confirmation and a full dataset for ACE-R/CBI-R (n=54) and PSP-RS (n=17) scores, analysis of covariance (ANCOVA) tested differences in ACE-R, CBI-R & PSP-RS subdomains across pathological groups, adjusting for age at assessment, sex, years of education and proportion of total symptom duration. Due to the high number of variables, ANCOVA p-values were adjusted for False Discovery Rate (FDR; using the Benjamini-Hochberg procedure), while post-hoc comparisons were corrected using Tukey’s HSD. 6 values for missing years of education were imputed using the MICE package.
2. **Dimensionality Reduction and Composite Scores:** A Principal component analysis (PCA; *FactoMineR* package) was applied to ACE-R and CBI-R domain scores across all patients who had complete data for both measures (n=122). Suitability for PCA was confirmed by Kaiser-Meyer-Olkin (KMO = 0.84) and Bartlett’s test of sphericity (p<<0.001). Components were retained based on Cattell’s scree criterion and eigenvalues > 1. A varimax rotation was performed on the retained components to improve interpretability. For patients who also had neuropathological examination (n=54), group differences across pathological groups in retained component scores were assessed using ANCOVA with the same covariates and pairwise comparisons with Tukey’s HSD correction.
3. **Predictive modelling of pathology:** For patients with full ACE-R & CBI-R scores and neuropathological examination (n=54), we used raw subdomain scores to predict final pathological diagnosis via Support Vector Machine (SVM; Radial Kernel, *e1071* package), Decision Tree, Random Forest (*randomForest* package), and Linear Discriminant Analysis (LDA; *MASS* package). Models were asked to predict primary AD pathology from other pathologies. Models employed a fixed seed (default 42) and 10-fold cross-validation with 30 repeats to ensure consistency of results. Within-resample up-sampling of the minority AD class was performed to mitigate class imbalance. Up-sampling was performed on training folds only, with untouched validation folds to reflect real world distribution. An 80:20 train-test split was explored for generalisability but not used for primary accuracy estimates due to large error margins in small test sets. The best models were confirmed by Nadeau-Bengio corrected pairwise resampled t-tests, with Bonferroni adjusted p-values. Feature importance was determined using *caret*’s *varImp* function which is model specific.
4. **Biomarker analysis:** For patients with biomarker data (plasma NFL, pTau217, Aβ42/Aβ40 ratio, n=52), average levels and distributions were assessed. ANCOVA assessed group differences in biomarkers among participants with both biomarker and pathological data, adjusting for demographic covariates (n=13). One value for years of education was imputed to allow the ANCOVA. Associations between biomarkers and clinical variables were examined using Pearson correlations (FDR-adjusted) and linear regression.

### Standard Protocol Approvals, Registrations, and Patient Consents

The Cambridge Research Ethics Committee approved the study under two separate acquisition protocols: 12/EE/0475; 07/Q0102/3. Participants provided written informed consent in accordance with the Declaration of Helsinki.

### Data availability

Anonymised derived data at subject level that support the findings of this study are available from the corresponding author without restriction. Detailed clinical data may be requested but are subject to restrictions and likely need a material transfer agreement to preserve participant confidentiality.

## Results

### Demographics and clinical characteristics

Of the 226 participants with CBS, 72 had a neuropathological diagnosis: Alzheimer’s Disease (AD, n=22), Corticobasal Degeneration (CBD, n= 20), Progressive Supranuclear Palsy (PSP, n= 19), and 11 with other less common pathologies (including Synucleinopathies (SYNUC) such as Multiple Systems Atrophy (MSA), other tauopathies, and TDP-43, Figure 1). Co-pathologies were commonly identified but were not co-dominant and were present at a lower-level or early Braak/Kovacs stage. Participants with PSP pathology were significantly older than those with AD (Tukey HSD p=0.002), but otherwise there were no other demographic differences between groups (Table 1). To visualise the cognitive and clinical profile of each pathology, mean scores are shown in Figure 2. To visualise the distribution of scores, boxplots for each domain by pathology are shown in Supplementary Figure 1.

**Figure 1.**
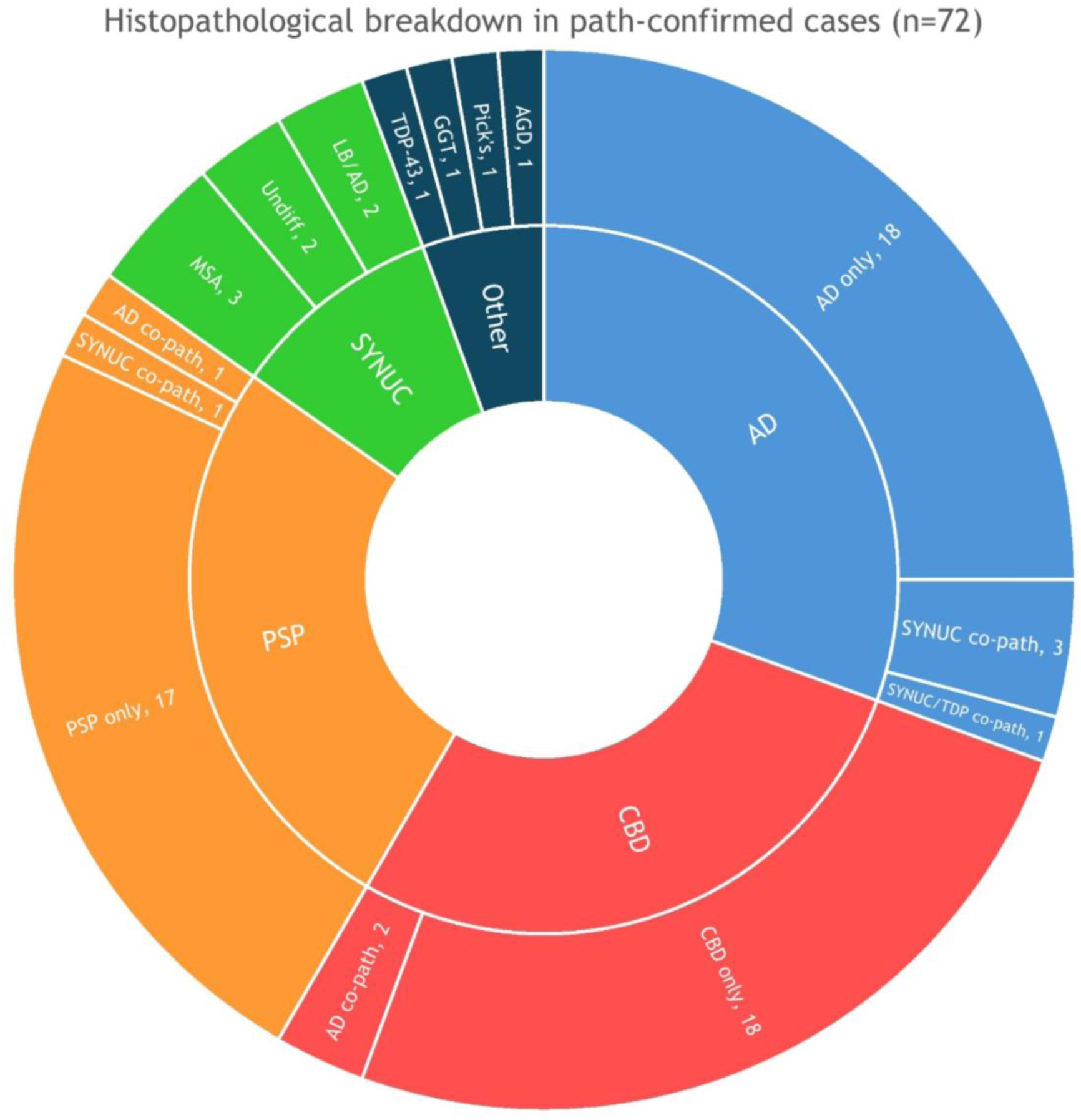
Histopathological breakdown of path-confirmed cases (n=72) with breakdown of co-pathology also shown. LB = Lewy Body pathology, SYNUC = Synucleinopathies, MSA = Multiple Systems Atrophy, Undiff = Undifferentiated Synucleinopathy, GGT = Globular Glial Tauopathy, Pick’s = Pick’s Disease, AGD = Argyrophilic Grain Disease, TDP-43 = TDP-43 pathology.

**Figure 2.**
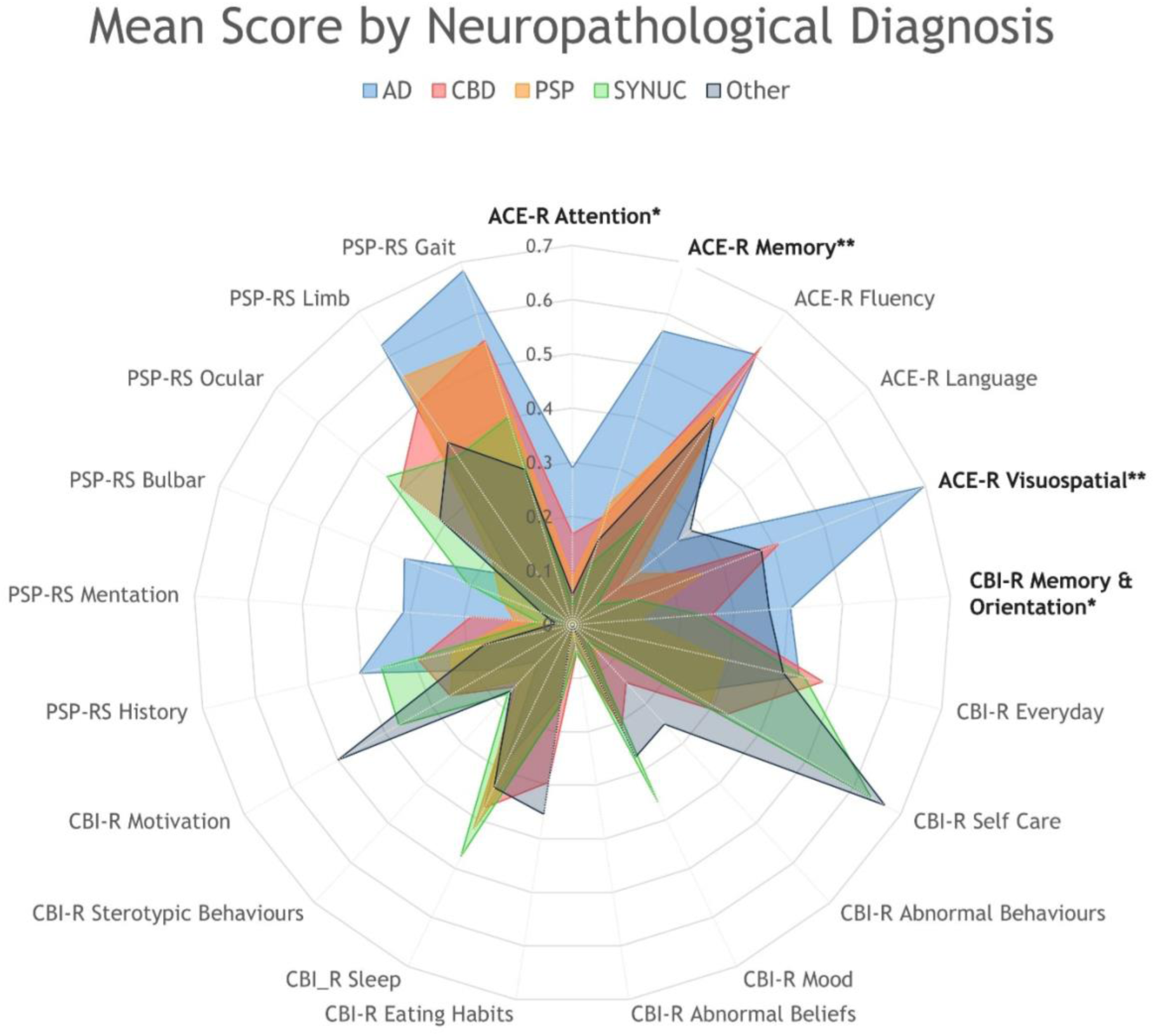
Radar Chart showing mean scores by histopathological diagnosis. For comparability, scores are given as a proportion of total score where a higher score corresponds to worse function. CBS-AD patients show worse function in ACE-R memory, ACE-R visuospatial, ACE-R attention and CBI-R memory & orientation. *ANCOVA FDR-adjusted p-value <0.05; ** FDR p<0.01

**Table 1.** Demographic table. All values are given in mean ± SD [range] for all 226 patients, and by pathology. Histopathological diagnoses are as in Figure 1. Total duration of symptoms was defined as either symptom onset to death, or symptom onset to current date if the patient is still alive. †PSP patients were significantly older than AD patients. *CBS-AD patients had significantly lower ACE-R total scores. ^a^FDR (False Discovery Rate Benjamini-Hochberg procedure) adjusted p value is shown from ANCOVA. ^b^ Significance for sex ratios were calculated separately with a Chi squared test.

| Variable | Total CBS | AD | CBD | PSP | SYNUC | Other | p-adj<sup>a</sup> |
| --- | --- | --- | --- | --- | --- | --- | --- |
| N | n=226 | n=22 | n=20 | n=19 | n=7 | n=4 | - |
| Age at first visit | 68.8 ± 8.7<br>[46.9–89.0] | 63.8 ± 8.5<br>[48.5–79.3] | 67.3 ± 6.5<br>[56.2–74.7] | **71.0 ± 3.6†**<br>[65.1–76.3] | 64.5 ± 7.7<br>[50.7–73.1] | 61.7 ± 7.8<br>[51.9–70.1] | **0.047†** |
| Sex (M:F) | 101:125 | 7:15 | 9:11 | 11:8 | 3:4 | 3:1 | ns ($\chi^2$)<sup>b</sup> |
| Symptom Duration at First Visit (Years) | 3.2 ± 2.2<br>[0.0–12.3] | 3.4 ± 1.9<br>[0.5–7.0] | 3.0 ± 1.5<br>[0.8–5.2] | 2.8 ± 2.7<br>[0.5–12.2] | 2.7 ± 0.8<br>[1.2–3.8] | 3.5 ± 1.7<br>[2.0–5.7] | ns |
| Total Symptom Duration (Years) | 7.9 ± 3.8<br>[0.3–29.8] | 9.3 ± 3.3<br>[4.2–14.9] | 7.6 ± 5.6<br>[1.9–29.8] | 6.2 ± 2.7<br>[2.8–14.3] | 6.5 ± 2.4<br>[4.0–11.2] | 10.7 ± 4.5<br>[5.3–15.7] | ns |
| Proportion of Total Symptom Duration at First Visit | 0.42 ± 0.21<br>[0.00–1.01] | 0.38 ± 0.21<br>[0.07–0.74] | 0.43 ± 0.18<br>[0.12–0.75] | 0.41 ± 0.22<br>[0.00–0.85] | 0.46 ± 0.19<br>[0.17–0.65] | 0.36 ± 0.19<br>[0.18–0.62] | ns |
| Years of Education | 12.1 ± 2.7<br>[5.0–20.0] | 12.1 ± 2.9<br>[9.0–18.0] | 11.8 ± 2.3<br>[9.0–16.0] | 11.2 ± 1.5<br>[10.0–15.0] | 13.4 ± 3.6<br>[10.0–19.0] | 13.0 ± 2.9<br>[10.0–16.0] | ns |
| ACE-R Total Score (max. 100) | 69.2 ± 21.1<br>[0.0–98.0] | **53.7 ± 22.2\***<br>[13.0–98.0] | 73.3 ± 16.7<br>[35.0–96.0] | 76.9 ± 16.4<br>[27.0–94.0] | 87.3 ± 9.6<br>[74.0–96.0] | 73.8 ± 18.7<br>[47.0–90.0] | **0.001\*** |
| CBI-R Total Score (max. 180) | 42.9 ± 30.8<br>[0.0–142.0] | 39.1 ± 26.1<br>[8.0–84.0] | 46.0 ± 37.7<br>[9.0–142.0] | 32.5 ± 17.4<br>[0.0–67.0] | 50.4 ± 25.4<br>[23.0–91.0] | 61.7 ± 25.4<br>[36.0–77.0] | ns |
| PSP-RS Total Score (max. 100) | 33.9 ± 17.0<br>[1.0–82.0] | 43.3 ± 17.1<br>[32.0–63.0] | 35.5 ± 30.4<br>[14.0–57.0] | 31.0 ± 11.9<br>[16.0–46.0] | 32.3 ± 2.1<br>[30.0–34.0] | 22.5 ± 9.2<br>[16.0–29.0] | ns |

#### Cognitive profile

ACE-R total scores were significantly lower in CBS-AD compared to other diagnoses (FDR p=0.002). Pairwise comparisons showed CBS-AD had worse scores than PSP (p=0.002) and SYNUC groups (p=0.012), with a trend toward lower scores than CBD (p=0.073) (Supplementary Table 1).

On subdomains, CBS-AD patients performed significantly worse with large effect sizes (Hedge’s g = 1.21-1.95, Supplementary Table 1) in:

- **ACE-R memory** (p=0.002; vs PSP p=0.005, vs CBD p=0.006, vs SYNUC p=0.050)
- **ACE-R visuospatial** (p=0.002; vs PSP p=0.008, vs SYNUC p=0.013)
- **ACE-R attention** (p=0.012; vs PSP p=0.026)
- **CBI-R memory & orientation** (p=0.028; vs PSP p=0.045)

Across all groups, ACE-R fluency was markedly impaired, and CBI-R scores indicated severe difficulties in everyday activities, self-care, and sleep, highlighting poor ADL functioning in CBS patients.

#### Motor function

PSP-RS scores did not differ significantly (noting that only 17 path-confirmed cases had PSP-RS scores). Nevertheless, gait and limb dysfunction were prominent across all pathologies, especially AD, CBD, and PSP. CBS-AD patients showed slightly worse bulbar, and mentation scores.

### Group differences in composite clinical scores

We conducted a PCA on CBS patients with complete ACE-R and CBI-R data (n = 122), including the 54 with confirmed pathology. PSP-RS domains were excluded due to missing data, and no imputation was performed. Scores were scaled to 0–1, and ACE-R domains were inverted so that higher values reflected greater impairment.

Three principal components were retained and subjected to varimax rotation (Figure 3A; Supplementary Figure 2):

- **PC1 (40.0% variance; eigenvalue 6.00):** behavioural dimension, with strong positive loadings across CBI-R behavioural domains.
- **PC2 (17.2% variance; eigenvalue 2.58):** amnestic cognitive dimension, defined by strong positive loadings on ACE-R (particularly attention and memory) and minimal contribution from CBI-R.
- **PC3 (7.3% variance; eigenvalue 1.09):** functional dimension, with high loadings on CBI-R everyday skills and self-care.

**Figure 3.**
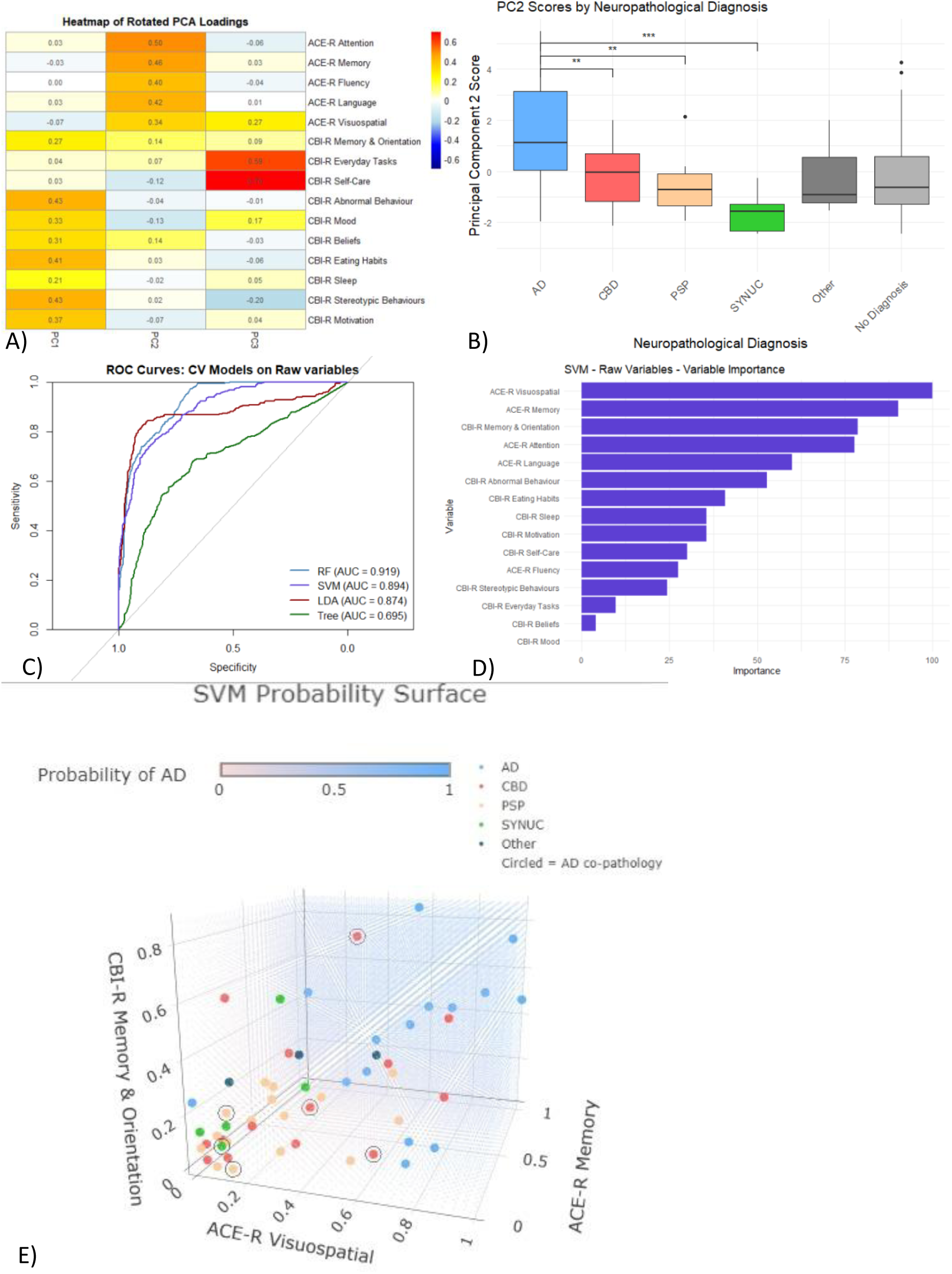
PCA and predictive modelling results. **(A**) Heatmap of loadings for the first three principal components: PC1 (behavioural; CBI-R), PC2 (cognitive; ACE-R), and PC3 (functional; CBI-R). **(B)** PC2 scores by pathology group, showing significantly higher (worse) scores in CBS-AD compared with CBD, PSP, and synucleinopathies. **(C**) ROC curves for the four classification models; Random Forest, SVM, and LDA show similarly high AUCs. **(D)** Variable importance for the SVM model, with ACE-R visuospatial, ACE-R memory, and CBI-R memory & orientation identified as the strongest predictors. **(E)** SVM probability surface illustrating predicted AD probability across these three key variables, overlaid with patient data points coloured by pathology; cases with secondary AD co-pathology are circled. Higher impairment across these domains corresponds to increased predicted probability of AD pathology.

ANCOVA on PC scores showed no group differences in PC1 or PC3. However, patients with CBS-AD had significantly higher PC2 scores (indicating worse cognition) than all other pathology groups (p < 0.001). Post-hoc comparisons confirmed poorer cognition in CBS-AD relative to CBD (p = 0.009), PSP (p = 0.005), and synucleinopathies (p = <0.001). Effect sizes were large (Hedge’s g = 1.42–2.28; Figure 3B; Supplementary Table 2).

### Predictive Modelling

We next evaluated machine-learning models for predicting AD pathology in CBS patients using raw ACE-R and CBI-R subdomain scores. Analyses were restricted to patients with confirmed pathology and complete clinical data (n = 54). Models included Decision Tree, LDA, Random Forest (RF), and SVM.

#### Model Performance

The RF classifier achieved the highest overall performance: **Accuracy:** 85.6% (95% CI: 83.8–87.3%); **Sensitivity:** 59.3%; **Specificity:** 95.7%; **AUC:** 0.919; **Brier score:** 0.120 (Figure 3C; Supplementary Table 3). SVM and LDA performed comparably, with accuracies of 84.6–85.6% and AUCs of 0.879–0.912. Confusion matrices and full performance metrics are provided in Supplementary Figure 3 and Supplementary Table 3.

Statistical comparison using paired resampled t-tests with the Nadeau–Bengio correction (and Bonferroni-adjusted p-values) showed no significant differences among SVM, RF, and LDA for either accuracy (p = 1.00 for all comparisons) or AUC (RF vs SVM p = 0.452; RF vs LDA p = 0.256; SVM vs LDA p = 1.00). The Decision Tree model performed significantly worse than the other three models across all metrics (p < 2×10⁻¹⁶).

All models demonstrated held-out test accuracy between 80–90%, indicating generalisability, though confidence intervals were wide due to the small test set. Calibration curves showed good calibration for SVM and RF, with minor under-confidence at low predicted AD probabilities and over-confidence at high probabilities for RF (Supplementary Figure 5).

#### Feature Importance

Across all models, the most influential predictors of AD pathology were ACE-R visuospatial, ACE-R memory, and CBI-R memory & orientation scores (Supplementary Figure 4). The SVM probability surface for these top predictors (Figure 3D–E) demonstrated that higher impairment in these domains corresponded to increased probability of AD pathology.

### Biomarker analysis across pathological groups

Plasma NFL, pTau217 and Aβ42/Aβ40 ratio were available for 52 patients (2013–2022). Pathological confirmation was available in 13/52 cases. ANCOVA revealed no significant differences in biomarker levels between AD and other pathologies (Supplementary Table 4).

To assess clinical relevance, we compared each patient’s biomarker values with established AD cut-offs (20,22,23,31) (Figure 4). All 52 patients fell below the Aβ42/Aβ40 ratio threshold (<0.12), which would classify every case as “AD-positive” despite the absence of AD pathology or co-pathology in many individuals. NFL levels exceeded the cut-off in 34/52 patients, and pTau217 levels were above the AD-positivity threshold in 17/52. Among the six patients with neuropathological data who met the pTau217 cut-off, only two had AD pathology, while the remaining four had neither primary AD nor AD co-pathology. Overall, biomarker positivity showed no consistent relationship with histopathological AD diagnosis, with most pTau217-positive patients lacking AD pathology.

**Figure 4.**
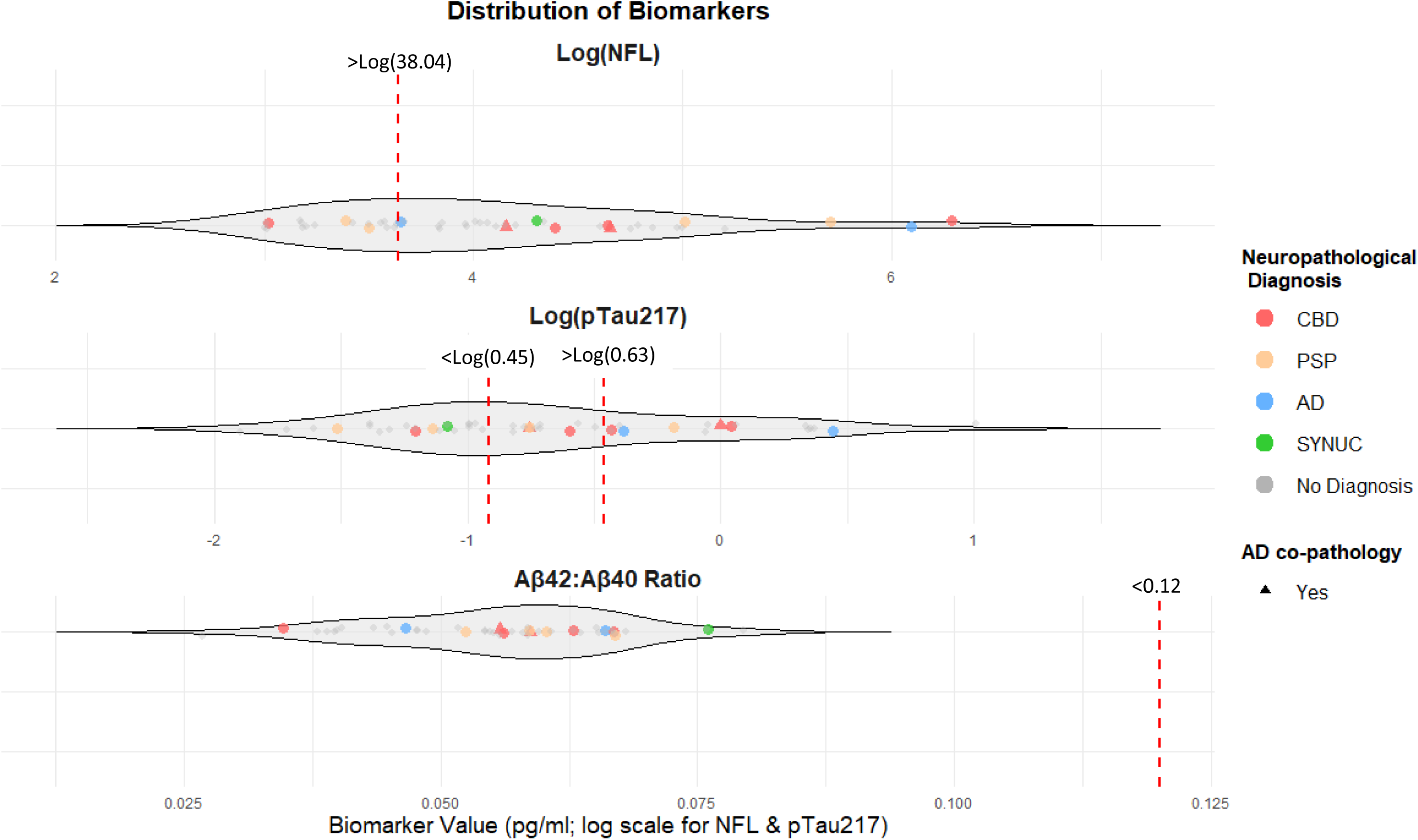
Plasma biomarker distributions. Violin plots show plasma NFL, pTau217, and Aβ42:Aβ40 ratio values (pg/mL). NFL and pTau217 are displayed on a logarithmic scale. Cases with neuropathological confirmation are colour-coded by diagnosis; those without post-mortem data are shown in grey. Individuals with a non-AD primary pathology and secondary AD co-pathology are marked with triangles. Published cut-offs for AD positivity are indicated by dotted lines (NFL > 38.04; pTau217 > 0.63 or < 0.45; Aβ42:Aβ40 < 0.12).

### Biomarker correlations with clinical scores

We assessed associations between plasma biomarkers and clinical, cognitive, and demographic variables using pairwise correlations and linear regression models. NFL and pTau217 values were log-transformed to improve normality.

Higher NFL levels were associated with poorer cognitive performance, including ACE-R attention (FDR-adjusted p = 0.019) and memory, fluency, and language domains (uncorrected p < 0.05). NFL was also associated with fewer years of education (uncorrected p < 0.05). Higher pTau217 levels correlated with lower scores on the stereotypic behaviour subscale of the CBI-R (uncorrected p < 0.05; Supplementary Figure 6A–B).

Linear regression models adjusting for disease duration yielded similar patterns but accounted for only a small proportion of variance (R² = 0.095–0.20; Supplementary Figure 6C). >Log(38.04)

## Discussion

The principal result of this study is that the cognitive profile of CBS differs according to the underlying pathology. Using PCA-derived composite scores and simple machine learning, the clinical cognitive profiles predicted Alzheimer’s disease pathology with high accuracy (∼85%, AUC 0.874-0.919), beyond what can currently be achieved with available blood-based biomarkers (25). Positive pTau217 did not accurately predict presence of AD pathology nor exclude CBD pathology, in the context of a corticobasal syndrome.

CBS-AD patients had significantly lower ACE-R total scores and worse memory and visuospatial performance, as well as poorer CBI-R memory and orientation scores at first visit. These findings align with prior work showing that people with path-confirmed CBD and PSP have worse verbal fluency and impaired ADLs, while people with synucleinopathies such as MSA have relatively preserved cognition, and people with AD have the most severe cognitive deficits (15,17,18,33–35). PCA revealed that CBS-AD patients cluster strongly on a global cognitive impairment dimension (PC2), reinforcing that widespread cognitive decline - not just focal deficits - distinguishes AD pathology. Prior studies using amyloid PET or CSF biomarkers have suggested similar trends (36–42), but these methods risk misclassification due to high amyloid positivity (up to 20-57% (43)) in non-AD tauopathies, and age-related positive PET in the absence of AD pathology, especially given previous evidence that 10-44% of people with normal cognition over the age of 50 show amyloid positivity by PET/CSF biomarkers (44). Our large effect sizes (Hedge’s g 1.0-2.0) confirm that memory and visuospatial impairment are robust indicators of AD pathology within CBS, consistent with previous small-scale studies and meta-analyses (18). Our use of histopathology as the gold standard and detailed cognitive scoring provides stronger evidence that global cognitive decline is a key marker of AD pathology in CBS. (4,5,7,9,11,12,14).

The Random Forest model achieved 85.6% accuracy and excellent calibration using only clinical cognitive scores, with visuospatial and memory scores as the strongest predictors. While few studies have explored predictive modelling for pathology within CBS (15,16), most prior work has focused on differentiating clinical syndromes or predicting conversion along the Alzheimer’s continuum using multimodal data, including imaging and CSF biomarkers (19–21). These approaches often achieve AUCs in the range of 0.87–0.96 when combining neuropsychological scores with imaging or biomarker data (19,20), but they require costly and invasive procedures that limit scalability. In contrast, we found that pen-and-paper cognitive assessments - widely available and inexpensive - provide highly accurate predictions of AD pathology within CBS, even in the absence of advanced imaging or CSF analysis. Such methods would be suitable in poor resourced settings.

This finding is particularly relevant given the poor clinicopathological correlation in CBS, where clinical diagnosis alone, and presence or absence of singular clinical features, often fail to predict underlying pathology (4–6,8,11,12,14). Previous studies using amyloid PET or CSF biomarkers to infer AD status in CBS have reported similar cognitive patterns to ours - worse memory and visuospatial function in CBS-AD - but these methods are not definitive and can misclassify patients due to high rates of amyloid co-pathology in non-AD tauopathies (43,44). Our use of histopathology as the gold standard strengthens the validity of these associations and supports the concept that detailed cognitive profiling can serve as a robust surrogate marker for pathology.

Importantly, the logic of our predictive model aligns with established clinical observations: CBS-AD patients typically exhibit widespread cognitive impairment, particularly in memory and visuospatial domains, whereas CBS-CBD and CBS-PSP patients show more focal deficits in executive function and fluency (15–18,33–35). This suggests that machine learning models trained on structured cognitive data are not only accurate but also clinically interpretable.

Surprisingly, plasma biomarkers did not reliably distinguish AD pathology within CBS. All patients fell below the commonly used Aβ42/Aβ40 ratio threshold for AD positivity (22,31), despite only ∼30% having AD pathology. This discrepancy likely reflects the ratio’s sensitivity to early amyloid deposition, which can precede dementia by years (25,31,44). While Aβ42/Aβ40 ratio performs well in cognitively normal or mildly impaired individuals (AUC ∼0.81–0.91) (24,25), its discriminative power diminishes in advanced disease and mixed-pathology syndromes, as seen in our cohort and others (24). Recent studies suggest that Aβ42/Aβ40 ratio may be most useful for screening preclinical AD or predicting future amyloidosis (31), rather than differentiating pathology someone with symptomatic CBS where multiple neurodegenerative processes may coexist. These findings highlight the risk of false positives when applying plasma biomarker thresholds in clinically heterogeneous CBS cohorts.

pTau217 is currently a leading blood biomarker for AD (AUC 0.91–0.96 in diverse large cohorts) (20,21,21,25,46,47), but did not achieve high specificity in our study of CBS. Several factors may explain this lower than expected performance of pTau217. First, our sample size for biomarker-pathology comparisons was small, limiting statistical power. Second, many validation studies for pTau217 have compared AD to healthy controls or MCI, not to non-AD dementias (20,25). This inflates specificity estimates, which may drop substantially for other group contrasts. Indeed, even where histopathology-based studies reporting strong performance for pTau217 (AUC ∼0.95) (21,47,48), co-pathology remains a major challenge. If treatment trials rely solely on blood-based biomarkers to determine AD status, there is a real risk of enrolling patients whose primary pathology is a 4R-tauopathy with secondary amyloid deposition, potentially undermining group-level therapeutic effects.

NFL, by contrast, showed negative correlations with cognitive performance but it remains a non-specific marker of axonal injury and neurodegeneration (23,49,50). Elevated NFL is common across atypical parkinsonian disorders and even higher in PSP and CBD than in AD (49,50), explaining its poor discriminative value in our cohort. While NFL may still have utility for differentiating atypical parkinsonism from PD, or predicting survival (49,51), it is unlikely to aid in pathology-specific diagnosis within CBS.

Taken together, these findings highlight the need for caution when applying blood-based biomarkers for diagnostic decision-making or trial enrolment in CBS. While biomarkers such as pTau217 hold promise for early detection and risk stratification, their specificity in mixed pathology remains uncertain. Our results suggest that detailed neurocognitive profiling - cheap, scalable, and clinically interpretable - should remain central to improving clinicopathological correlation, with biomarkers serving as complementary tools rather than standalone determinants.

There are limitations to our study. First, our cohort, while larger than most prior clinico-pathological studies, remains relatively small and retrospective. Second, we did not have a clinical rating scale for movement disorders (e.g: UPDRS or PSP-RS) for many participants, which limits the power to detect correlations with motor severity. Third, we did not have CSF biomarker results and only had plasma biomarkers on a subset of participants at a single time point. We also did not have access to levels of the recently described Microtubule Binding Region (MTBR) biomarkers for AD (such as MTBR-tau243), which may be more specific for tau tangle pathology (55). Our predictive modelling approach has inherent limitations. The sample size and class imbalance increase the risk of overfitting, as reflected by wide confidence intervals on held-out data testing. Additionally, PCA-based feature reduction may capture cohort-specific variance, and the use of single time-point data prevents assessment of longitudinal predictive power. Finally, while our predictive models achieved high accuracy, their complexity reduces interpretability compared to simpler models.

In conclusion, this study highlights that detailed neurocognitive profiling can accurately predict AD pathology within CBS, offering a practical alternative to biomarker-based approaches. While plasma biomarkers such as pTau217 show promise, their specificity was limited. Our findings highlight the continued importance of structured cognitive assessment as a scalable, low-cost tool to improve diagnostic precision and guide patient selection for targeted therapies.

## Acknowledgements

We thank our patients, their carers, clinicians, research nurses, research assistants; and all other staff at the Cambridge Biomedical Research Centre and Centre for Parkinson-plus.

## Author contributions

NH and JBR contributed to the conception and design of the study. SS and NH were involved in data acquisition, analysis, drafting the manuscript, and prepared the figures and tables. AE, RC, AM, GS, KS, MR, MN, MM, TR, JBR contributed to data acquisition (neuropathology AQ, KA), and edited the final version of the manuscript.

## Financial Disclosures of all authors (for the preceding 12 months)

The study was supported by the Wellcome Trust (220258), the Cambridge Centre for Parkinson-Plus (RG95450); the NIHR Cambridge Biomedical Research Centre (NIHR203312); Medical Research Council (MC_UU_00030/14; MR/T033371/1); Dementias Platform UK (RG94383; and G103658); Association of British Neurologists, Patrick Berthoud Charitable Trust (RG99368), PSP Association (G132180), the Academy of Medical Sciences (G130967), Rosetrees-Race Against Dementia Teams Award (G124579), and Race Against Dementia Alzheimer’s Research UK (ARUK-RADF2021A-010). The views expressed are those of the authors and not necessarily those of the NIHR or the Department of Health and Social Care. For the purpose of open access, the author has applied a CCBY public copyright licence to any Author Accepted Manuscript version arising from this submission.

## Notes

**Financial Disclosures/conflict of interest:** The authors do not have any competing interest pertaining to this manuscript.

### Competing Interest Statement

The authors have declared no competing interest.

