## Supplementary for "Cognitive profile accurately predicts underlying pathology in Corticobasal Syndrome"

### Supplementary Materials

| Variable | ANCOVA | | Tukey’s HSD | | Effect size (Hedge’s g [95% CI]) |
| --- | --- | --- | --- | --- | --- |
|  | F Value | p-adj | Mean Diff | p-value |  |
| ACE-R Total  AD vs SYNUC  AD vs PSP | 6.73 | 0.002 | 0.284  0.261 | 0.012  0.002 | 2.19 [0.83-3.55]  1.67 [0.55-2.80] |
| ACE-R Memory  AD vs SYNUC  AD vs PSP  AD vs CBD | 6.58 | 0.002 | 0.375  0.359  0.311 | 0.050  0.005  0.006 | 1.84 [0.32-3.36]  1.45 [0.19-2.71]  1.70 [0.67-2.72] |
| ACE-R Visuospatial  AD vs SYNUC  AD vs PSP | 6.55 | 0.002 | 0.569  0.442 | 0.013  0.008 | 1.95 [0.22-2.73]  1.47 [0.22-2.73] |
| ACE-R Attention  AD vs PSP | 4.76 | 0.012 | 0.228 | 0.026 | 1.47 [0.21-2.72] |
| CBI-R Memory & Orientation  AD vs PSP | 4.11 | 0.022 | 0.283 | 0.045 | 1.21 [-0.04-2.47] |
| ACE-R Language  Other vs CBD | 3.77 | 0.028 | 0.235 | 0.046 | 2.86 [0.95-4.78] |

**Supplementary Table 1 – Significant ANCOVA and Tukey HSD results for raw clinical variables.** All variables showing significant ANCOVA effects are listed with F‑values and FDR‑adjusted p‑values (Benjamini–Hochberg method). For each significant result, Tukey HSD pairwise comparisons are provided, including mean differences and associated p‑values. Scores are expressed as proportions of the total domain score, where higher values indicate greater impairment. AD patients demonstrated significantly worse performance than CBD, PSP, or synucleinopathy groups in ACE‑R memory, visuospatial, and attention domains, total ACE‑R score, and the CBI‑R memory & orientation domain.

| Principal Component | ANCOVA | |  | Tukey’s HSD | | Effect size (Hedge’s g [95% CI]) |
| --- | --- | --- | --- | --- | --- | --- |
|  | F Value | p-value | Comment | Mean Diff | p-value |  |
| PC1 | 0.87 | 0.492 | Not significant |  |  |  |
| PC2  AD vs SYNUC  AD vs PSP  AD vs CBD | 6.75 | <0.001 | *Significant* | 3.20  2.32  1.99 | <0.001  0.005  0.009 | 2.28 [0.92-3.64]  1.65 [0.53-2.78]  1.42 [0.39-2.45] |
| PC3 | 1.41 | 0.247 | Not significant |  |  |  |

**Supplementary Table 2 – ANCOVA and Tukey HSD results for PCA components.** ANCOVA results for PC1–PC3 are presented with F‑values and raw p-values. For significant effects, pairwise comparisons are provided, including mean differences, with Tukey HSD corrected p-values. As reported in the main text, AD patients show significantly higher PC2 scores (greater cognitive impairment) than those with CBD, PSP, or synucleinopathy.

| Model | 10-fold Cross Validation, 30 repeats | | | | | | Held-out Testing (80:20 split) | | |
| --- | --- | --- | --- | --- | --- | --- | --- | --- | --- |
|  | Acc. (%)  [95% CI] | Sens. (%) | Spec. (%) | Bal Acc. (%) | AUC | Brier | Acc. (%) [95% CI] | Sens. (%) | Spec. (%) |
| SVM | 84.6  [82.7-86.3] | 63.5 | 92.7 | 78.1 | 0.894 | 0.115 | 90.0  [55.5-99.8] | 100.0 | 85.7 |
| Random Forest | 85.6  [83.8-87.3] | 59.3 | 95.7 | 77.5 | 0.919 | 0.120 | 80.0  [44.4-97.5] | 33.3 | 100.0 |
| LDA | 84.9  [83.0-86.6] | 84.7 | 85.0 | 84.8 | 0.874 | 0.123 | 80.0  [44.4-97.5] | 100.0 | 71.4 |
| Decision Tree | 71.0  [68.7-73.2] | 58.9 | 75.6 | 67.3 | 0.695 | 0.223 | 80.0  [44.4-97.5] | 66.7 | 85.7 |

Supplementary Table 3 – Predictive modelling performance metrics. Raw accuracy, sensitivity, specificity, balanced accuracy, AUC, and Brier scores are reported for each model (SVM, Random Forest, LDA, and Decision Tree) trained on raw clinical variables. Ninety‑five percent confidence intervals for raw accuracy are provided in brackets. Held‑out test performance is also shown. SVM, Random Forest, and LDA models demonstrated similarly high accuracy.

| Variable | Total CBS | AD | CBD | PSP | SYNUC | ANCOVA | |
| --- | --- | --- | --- | --- | --- | --- | --- |
|  |  |  |  |  |  | F Value | p-value |
| Biomarker Data Available (N)  **AD co-pathology | n=52 | n=2 | n=6    n=2 | n=4    n=4 | n=1 |  |  |
| NFL (pg/ml) | 81 ± 97  [20–539] | 241 ± 287  [39–444] | 152 ± 192  [21–539] | 129 ± 128 [30–302] | 74.0 | 1.51 | 0.321 |
| pTau217 (pg/ml) | 0.64 ± 0.55 [0.15–2.74] | 1.12 ± 0.62 [0.68–1.56] | 0.67 ± 0.30 [0.30–1.04] | 0.46 ± 0.27 [0.22–0.83] | 0.34 | 1.67 | 0.288 |
| Aβ42:Aβ40 ratio | 0.056 ± 0.01 [0.03–0.08] | 0.056 ± 0.01  [0.05–0.07] | 0.056 ± 0.01  [0.04–0.07] | 0.056 ± 0.01  [0.05–0.07] | 0.076 | 0.62 | 0.633 |

Supplementary Table 4 – Biomarker summary statistics by pathology. Mean ± SD and ranges for each biomarker are shown for the total CBS cohort and for each neuropathological group, along with the number of patients with available biomarker data. The number of CBD and PSP cases with AD co‑pathology is indicated. ANCOVA results (F‑values and p‑values) are provided in the final columns. No significant differences in biomarker levels were observed between neuropathological groups.

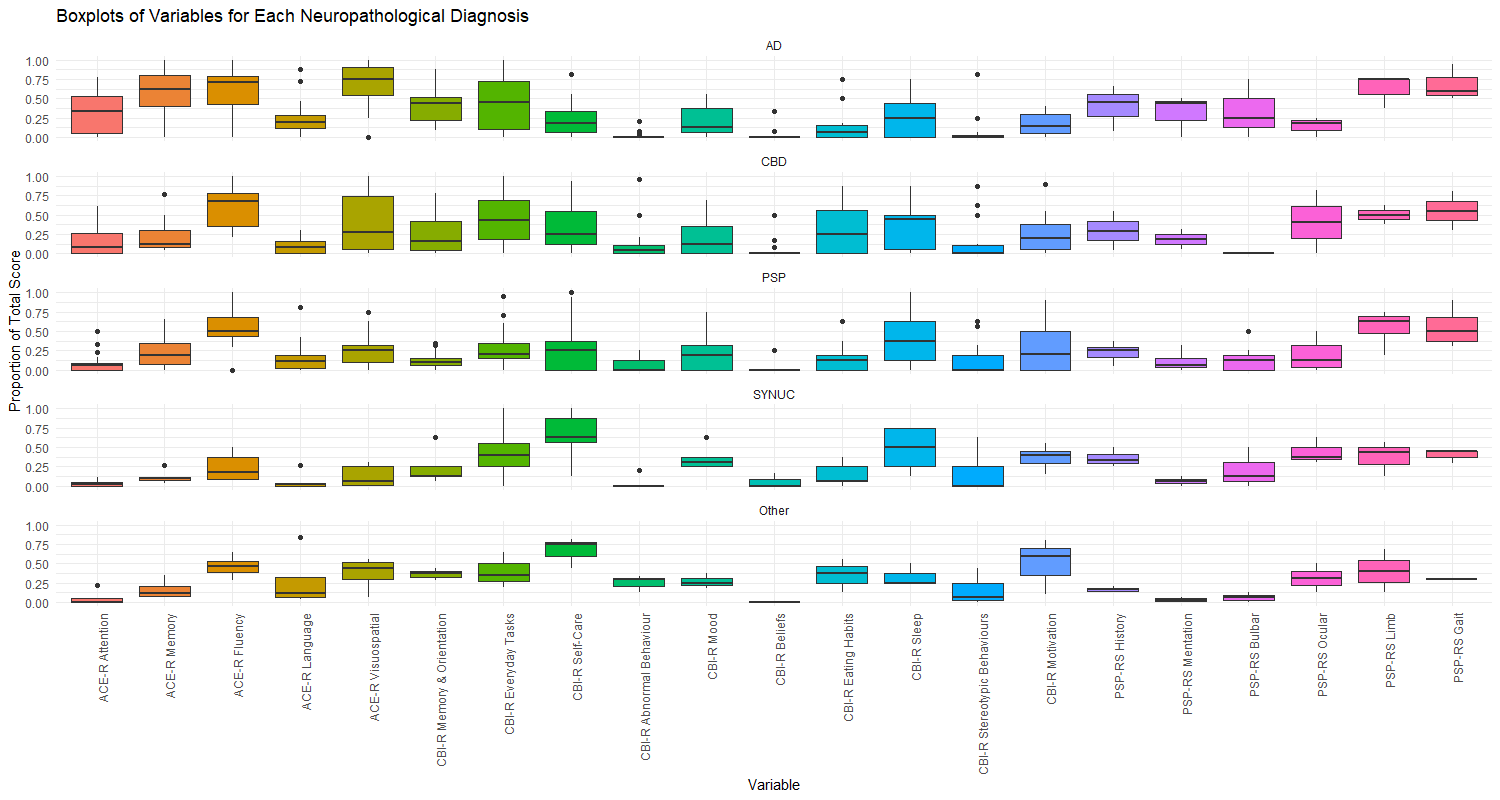

**Supplementary Figure 1.** Domain‑level clinical scores by pathology. Boxplots show median scores and interquartile ranges for each ACE‑R, CBI‑R, and PSP‑RS domain across histopathological groups. All scores are expressed as the proportion of the domain maximum; ACE‑R domains are presented as “points lost,” such that higher values reflect worse performance. Patients with AD pathology exhibit greater impairment in ACE‑R memory, ACE‑R visuospatial function, and CBI‑R memory & orientation.

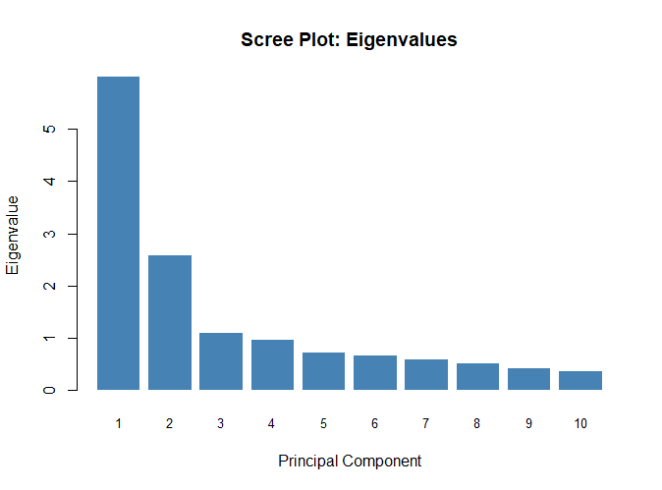

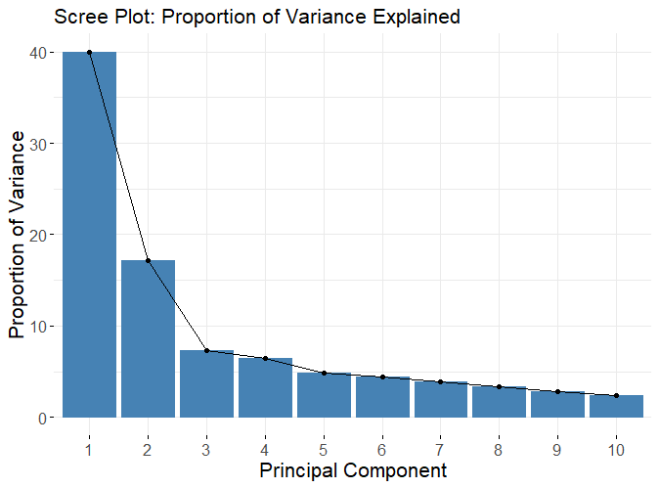

B)

A)

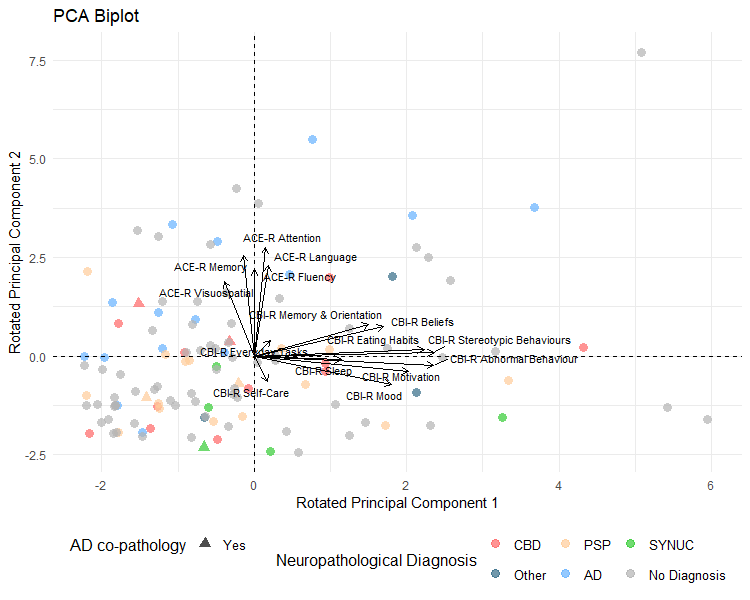

Supplementary Figure 2 – PCA results. (A) Scree plot of eigenvalues and (B) proportion of variance explained. The elbow occurs at PC3, and based on Cattell’s criterion and eigenvalues > 1.0, PC1–3 were retained for varimax rotation. (C) PCA biplot showing PC1 versus PC2 after rotation, with participant data points colour‑coded by diagnosis; cases with non‑AD primary pathology and AD co‑pathology are marked with triangles. Loading vectors for individual variables are displayed. PC1 reflects a CBI‑R behavioural dimension, whereas PC2 represents an ACE‑R cognitive dimension, with AD cases showing higher (worse) PC2 scores than other pathological groups.

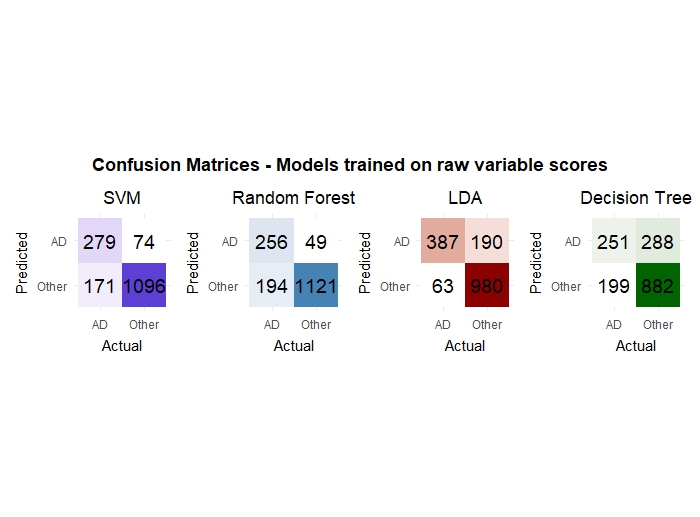

Supplementary Figure 3 – Confusion matrices for all predictive models. Confusion matrices for models trained on raw variable scores. The SVM, LDA and Random Forest models perform with similar accuracy, but the LDA model is more balanced between sensitivity and specificity.

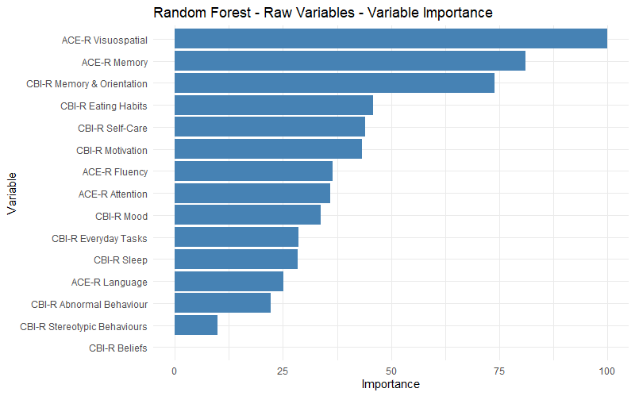

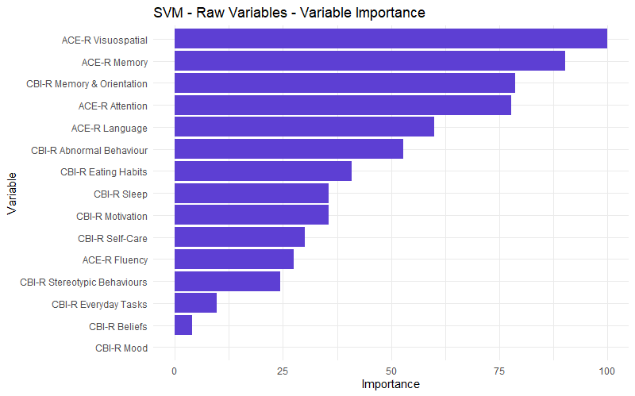

B)

A)

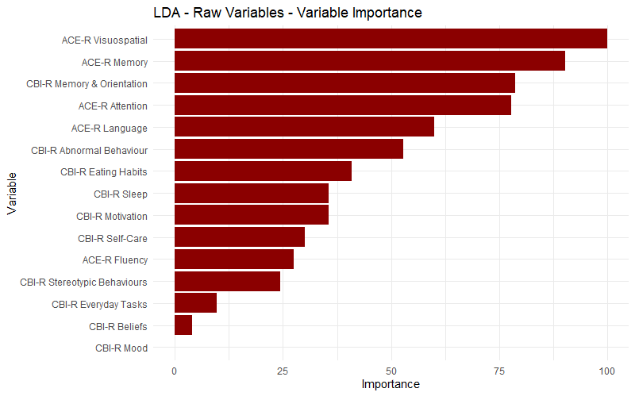

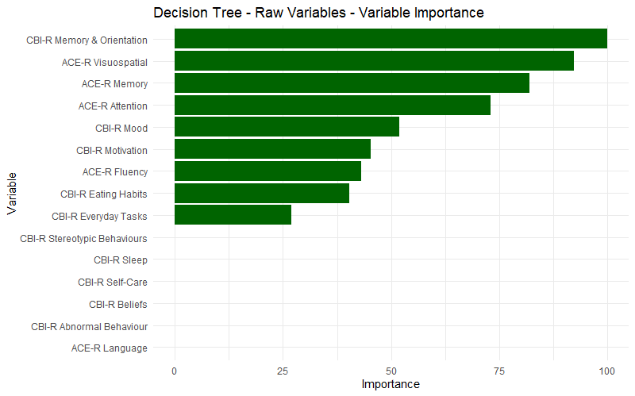

D)

C)

Supplementary Figure 4 – Charts showing variable importance for all predictive models for A) Random Forest B) SVM C) LDA D) Decision Tree. ACE-R visuospatial, memory and CBI-R memory & orientation are the most important variables across all models.
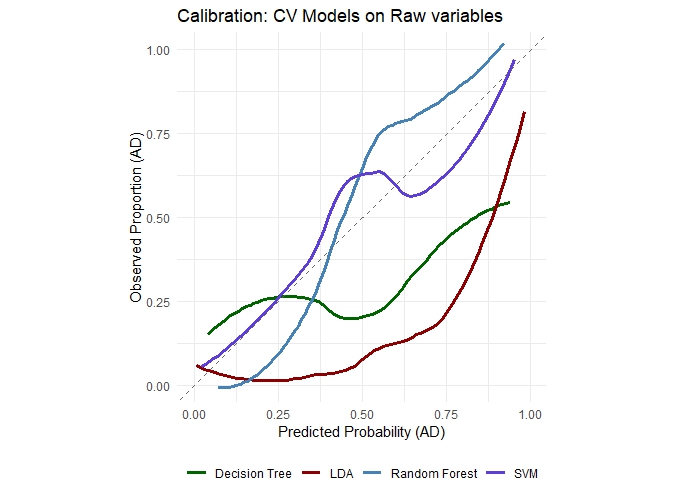

Supplementary Figure 5 – Calibration plots for predictive models. Calibration curves for all models trained on raw clinical variables are shown. The Random Forest and SVM classifiers demonstrate the best overall calibration.

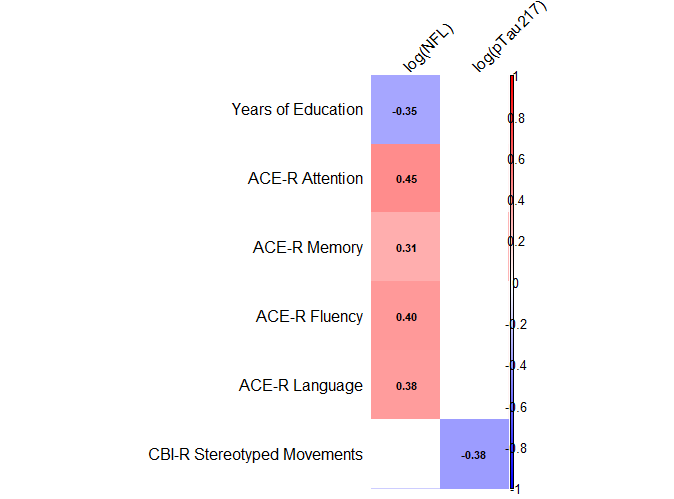

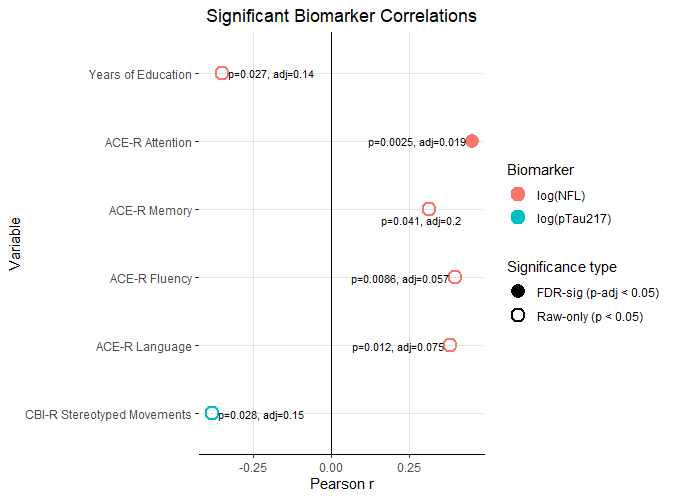

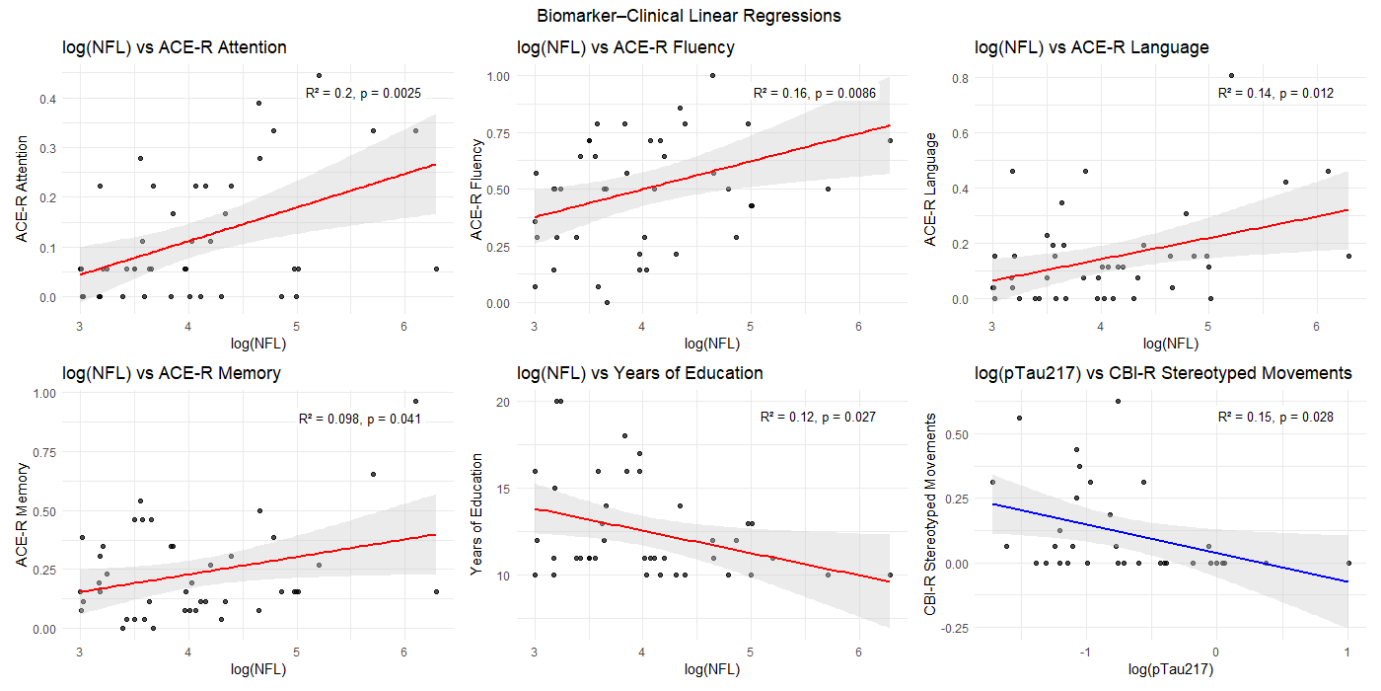

C)

B)

A)

Supplementary Figure 6 – (A) Heatmap of significant Pearson correlations between plasma biomarkers and clinical/demographic variables. Positive correlations are shown in red and negative correlations in blue. Log(NFL) is positively associated with ACE‑R cognitive impairment and negatively associated with years of education. Log(pTau217) shows a negative correlation with CBI‑R stereotyped movements. The Aβ42:Aβ40 ratio shows no significant correlations. (B) Dot plot illustrating the magnitude of Pearson correlation coefficients. Raw and FDR‑adjusted p‑values are shown; filled points indicate FDR‑significant correlations, and open points those significant only at the raw p‑value level. Only the correlation between Log(NFL) and ACE‑R attention remained significant after FDR correction. (C) Linear regression plots for the six significant associations, with fitted lines and 95% confidence intervals. Corresponding R² and p‑values are annotated. R² values are modest, indicating that biomarker levels explain only a small to moderate proportion of variance in these clinical measures.
